# Umbilical cord blood pTau217 and BD-tau are associated with markers of neonatal hypoxia: a prospective cohort study

**DOI:** 10.1101/2024.12.20.24319360

**Authors:** Emma Payne, Fernando Gonzalez-Ortiz, Kaitlin Kramer, Thomas Payne, Shreeya Marathe, Neha Mahajan, Ashly Liu, Jessica Barry, Andrew Duckworth, Mitchell Brookes, Bradley de Vries, Benjamin Moran, Helen Manning, Adrienne Gordon, Kaj Blennow, Henrik Zetterberg, David Zalcberg, Robert D. Sanders

**Author notes:** Corresponding authors: Emma Payne, David Zalcberg, address: St George Hospital, Gray St, Kogarah, NSW, 2217.

## Abstract

**Objective:** Current methods for early detection of hypoxic–ischemic encephalopathy (HIE) are limited by lack of specificity, cost, and time constraints. Blood tau protein concentrations reflect neuropathology in adults. This study examines tau as a potential HIE biomarker in neonates by relating cord blood levels to short-term fetomaternal outcomes. We aimed to examine 1) association of BD-tau with non-reassuring fetal status; 2) correlations between cord blood tau and other hypoxia biomarkers; 3) associations between tau levels and risk factors for fetomaternal morbidity; 4) associations between tau levels and short-term fetomaternal outcome.

**Methods:** 107 maternal participants were prospectively recruited at Royal Prince Alfred Hospital—a large Australian tertiary referral centre. Simoa analysis detected umbilical cord blood pTau217 and brain-derived (BD)-tau levels.

**Results:** Of 509 deliveries, cord blood was analysed in 107/110 recruited maternal participants. BD-tau correlated with non-reassuring fetal status (OR=3.0;95%CI=1.6– 5.7;p=0.001), though not when adjusting for mode of delivery and gestational age. BD-tau was higher in vaginal deliveries, and positively associated with pTau217, NfL, and lactate (p<0.001), and negatively associated with pH and base excess. pTau217 was higher in preterm neonates and was associated with neurofilament light chain (Spearman’s rho=0.44,p<0.001). BD-tau and pTau217 were associated with maternal hypertension and placental abnormalities.

**Conclusions:** Cord blood BD-tau correlates with surrogate markers of fetal hypoxia, whilst pTau217 may represent a marker of neurodevelopment. Further studies could explore whether these findings translate to clinical use of tau as an HIE biomarker.

**Funding:** US National Institutes of Health (grant:R01AG063849-01).

## Introduction

Hypoxic–ischemic encephalopathy (HIE)—a subtype of neonatal encephalopathy—is a syndrome of central nervous system (CNS) dysfunction caused by abnormalities in cerebral blood flow and impaired gas exchange perinatally, resulting in multiple organ failure.^1^ Approximately 40–60% of affected infants face mortality or severe neurodevelopmental disability by two years of age.^2^

Risk factors for HIE can broadly be categorised into antepartum (including fetal and maternal characteristics) and intrapartum factors. Understanding antepartum risk factors can complement intrapartum monitoring techniques used to detect ‘non-reassuring fetal status’ as defined by pathological fetal heart rate traces and fetal scalp blood sampling. Together, this information can identify fetuses at risk of hypoxic injury and guide decision-making regarding the urgency of delivery. However, intrapartum monitoring techniques are limited by a lack of specificity for pathological neurological damage, measurement difficulties, and poor inter-user reliability.^3,4^

The only proven neuroprotective treatment for HIE is therapeutic hypothermia (TH), which has only been validated in term or late preterm neonates.^2,5^ Severity of HIE can be classified as mild, moderate, or severe based on clinical signs and grading systems such as the modified Sarnat score.^6^ However, these scoring criteria are subjective, and grading may fail to identify neonates at risk of long-term adverse neurological sequelae; research has demonstrated abnormal neurological outcome in over 22% of cohorts of mild HIE.^5^ At this stage, TH is validated only in moderate to severe HIE, and there is insufficient evidence to recommend TH for mild HIE cohorts.^5^ Cord blood biomarkers may be helpful in identifying subgroups of mild HIE who would benefit from TH. Further aids to establish aetiology and severity of HIE include electroencephalogram (EEG) monitoring, and neuroimaging studies including MRI and cranial ultrasound.^3,7^ These tools are limited by time and cost constraints, require expert interpretation, and may be difficult to access in emergency situations.^7^ This indicates an ongoing need for objective, cost-effective, and rapid measures of identifying HIE in order to appropriately identify subgroups of HIE patients that could benefit from early intervention.

Real-time physiological biomarkers could address this gap in diagnosis and prognostication by providing objective point-of-care testing. Current cord or whole blood markers such as pH, lactate, and base excess remain non-specific for neurological injury.^2^ Tau is a promising biomarker of poor neurological outcome in adult cohorts with acute and chronic neurological disease.^8,9^ Studies of tau in neonates has, to date, focused on correlation of total blood tau and cerebral injury-related outcome measures. However, compared to total blood tau, specific tau subtypes—in particular pTau217 and ‘brain-derived tau’ (BD-tau)— have demonstrated greater specificity for adult neurological injury.^10,11^ This is the first study to examine tau subtypes in a neonatal cohort.

Given the paucity of evidence addressing tau subtypes and fetomaternal outcome, we conducted a prospective cohort study with the following aims:

1. Address feasibility of neonatal cord blood tau level measurement in an Australian cohort.
2. Assess the correlation between BD-tau and non-reassuring fetal status.
3. Assess correlations between cord blood tau—BD-tau, pTau217—and other cord blood biomarkers, maternal and neonatal risk factors for encephalopathy, and adverse outcome.

## Materials and Methods

### Study Population and Design

We performed a prospective cohort study of maternal and neonatal patients at the Royal Prince Alfred Hospital (RPAH), a tertiary referral centre in Sydney, Australia. Maternal participants of any age who were planned to birth at RPAH were eligible for study recruitment. Recruitment occurred prospectively from October 2022 at any time from 4 weeks prior to estimated date of delivery to the immediate post-partum period (with the option of retrospective consent). Trained recruiters obtained written, informed consent in English for all maternal participants in antenatal visits or on the birthing unit. Participants were excluded if they were non-English speaking, or had a history of psychological illness or other conditions that may interfere with capacity to provide informed consent after appropriate counselling in English.

Recruitment was undertaken as part of the BABBies (Benefits of Analysing Brain Biomarkers in perinatal care) Study—a prospective observational cohort study exploring the biomarkers neurofilament light chain (NfL), BD-tau, and pTau217. Data were stored in Sydney Local Health District (SLHD) REDCap electronic database. Ethics approval was obtained from SLHD Research and Ethics Committee (RPAH zone) (approval number: 2022/ETH01100). Our study was performed according to the National Statement on Ethical Conduct in Human Research (2007) and the CPMP/ICH Note for Guidance on Good Clinical Practice. Data reporting adhered to Strengthening the Reporting of Observational Studies in Epidemiology (STROBE) guidelines.

### Cord Blood Collection and Analysis

Umbilical venous cord blood was collected by the attending midwife in the immediate postpartum period. 1–3mL samples were collected and centrifuged in the RPAH Department of Anaesthetics laboratory, and plasma samples were then stored in deidentified cryovials.

Tau was measured on the Simoa HD-X platform with two-fold dilution factor in plasma. Plasma pTau217 and BD-tau were measured with previously validated assays.^12,13^ Signal variations within and between analytical runs were assessed using three internal quality control samples at the beginning and the end of each run.

### Outcome Data Collection

Demographic and outcome data were collected from RPAH electronic and paper medical records. Mode of delivery was classified categorically: vaginal delivery, emergency caesarean section, and elective caesarean section. Placental abnormalities were defined as any abnormality detected on placental ultrasound, on examination in the birth unit or in the operating theatre, or histopathology. The composite term ‘histopathological placental abnormality’ refers to only abnormalities detected on anatomical or histopathological examination per the 2014 Amsterdam Working Group nosology for classification of placental clinicopathological disorders.^14^ ‘Non-reassuring fetal status’ was defined as fetal blood sampling showing a pH ≤7.20 or lactate >4.7,^15,16^ or intrapartum cardiotocography (CTG) changes identified as ‘red zone’ criteria according to NSW Health electronic fetal monitoring guidelines (Table S1).^17^ ‘Non-reassuring fetal status’ did not include participants incidentally found to have abnormal umbilical cord blood values without meeting other criteria for non-reassuring fetal status.

### Outcomes

Our primary outcome was the association of BD-tau with non-reassuring fetal status. We adjusted this analysis for gestational age and mode of delivery, given these variables have previously been associated with both non-reassuring fetal status and cord blood concentrations of other biomarkers of acidosis and anaerobic metabolism.^18^ Secondary outcomes included the association of BD-tau and pTau217 with antenatal factors (gestational age, head circumference, birthweight), other cord blood biomarkers, maternal factors, delivery factors, and perinatal outcome. Resuscitation at birth was defined as cardiorespiratory support required following drying, warming, and mechanical stimulation.^19^

### Power Analysis

Our power analysis was based on the association of NfL with non-reassuring fetal status, which is reported separately.^20^ We used the previously reported baseline incidence of non-reassuring fetal status of 16%.^15^ Based on a two-sided t-test with α=0.025, a sample size of 110 participants provides 80% power to detect a Cohen’s d of 0.8 (i.e., a large effect size). The power analysis also applies to the primary outcome of this study, with a preserved type 1 error rate of α=0.05, as this value was halved a priori for the power calculation to account for additional analysis.

### Statistical Methods

#### Primary outcome

Both BD-tau and pTau217 concentrations showed a strong positive skew, hence were log_10_-transformed for all analyses. To determine the association of BD-tau with non-reassuring fetal status after adjusting for other variables, we used logistic regression (binomial family with logit link) with maximum likelihood estimation of the model parameters. The parametric G-formula was used to compute the mean risk difference from logistic regression models.^21^ We included a BD-tau*mode of delivery interaction in our model, as the relationship between neuronal biomarkers and fetal outcome would likely vary by birth route. To provide interpretable effect measures, we calculated the risk difference for the outcome per increase in BD-tau by its interquartile range (Q3 – Q1). The relative association of cord biomarkers with non-reassuring fetal status was quantified using the area under the receiver operator curve (AUROC). The 95% confidence interval for the AUROC was calculated using 2000 stratified bootstrap replicates.

#### Secondary outcomes

Bivariate biomarker correlations were assessed using rank-based nonparametric methods. The relative strength of biomarker correlations was tested using the method described by Meng, Rosenthal, and Rubin.^22^ For continuous secondary outcomes, we used linear regression with the ordinary least squares estimator for the model parameters. Associations between biomarker concentrations and placental abnormalities were adjusted for low birth weight (defined as birth weight <2.5kg, per World Health Organisation definition).^23^ Binary secondary outcomes were tested for linearity, and analysed in the same fashion as our primary outcome.

We used a p-value <0.05 to denote statistical significance. No adjustments for multiple comparisons were made in this preliminary study. All analyses were conducted in R using RStudio (Version 2024.04.0; R Foundation for Statistical Computing, Vienna, Austria). The ‘stats’ package was used for linear and generalised linear models. The ‘cocor’ package was used to compare the strength of bivariate biomarker concentrations.^24^ The ‘plotROC’ and ‘pROC’ packages were used for calculation and plotting of the AUROC.^25^

## Results

Over the study period 24^th^ October 2022 to 9^th^ December 2022, 110 maternal study participants were recruited from a total of 509 deliveries at RPAH. Cord blood was collected for a total of 108 participants, of whom 107 had either BD-tau or pTau217 levels available; 105 had BD-tau values and 106 had pTau217 values (STROBE diagram: Figure S1). Cohort demographic information is summarised in Table S2.

### Primary outcome

Increased BD-tau was associated with a higher risk of non-reassuring fetal status; each IQR increase in BD-tau was associated with an increased odds of fetal distress (OR=3.0; 95% CI:1.6, 5.7; p=0.001). However, this relationship was no longer observed when adjusting for mode of delivery and gestational age (OR=1.1; 95%CI=0.6, 1.2; p=0.667) (Table S3). BD-tau was not associated with non-reassuring fetal status when stratifying within each individual mode of delivery (Table S3). Comparative performances of BD-tau pH, lactate, and base excess in predicting non-reassuring fetal status are displayed in Figure 1. There was no association demonstrated between pTau217 and non-reassuring fetal status (Wilcoxon p=0.62).

**Figure 1:**
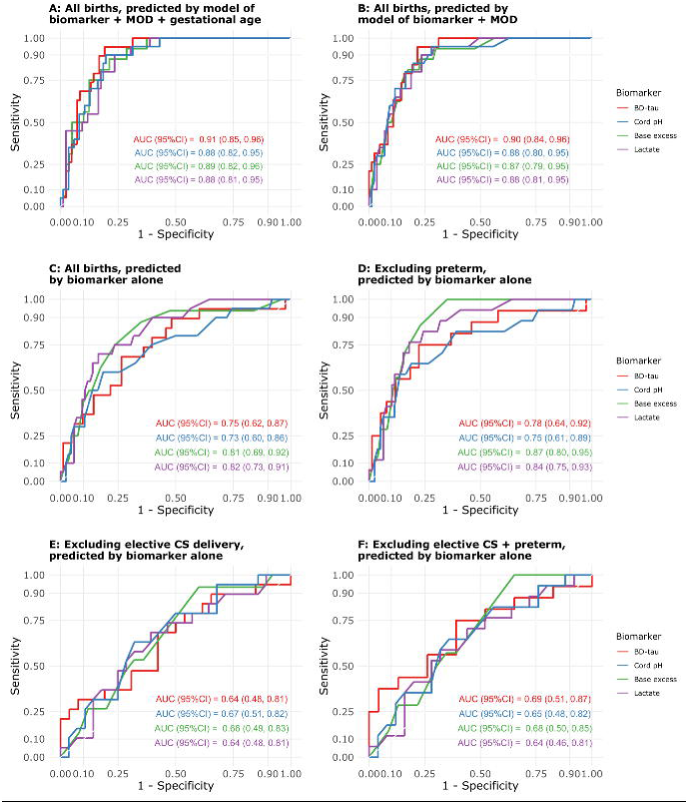
Receiver–operator curve (ROC) analysis of BD-tau, pH, base excess, and lactate in predicting binary non-reassuring fetal status. (A) ROC for the generalised linear model predicting non-reassuring fetal status and including the regressors: biomarker, mode of delivery, and gestational age. (B) ROC for the generalised linear model predicting non-reassuring fetal status and including the regressors: biomarker and mode of delivery. ROC for the biomarkers only, including (C) all births, (D) excluding preterm births, (E) excluding elective caesarean section, and (F) excluding elective CS and preterm birth.

### Secondary outcomes

#### Antenatal Factors

##### Relationship between tau and fetal factors

pTau217 was negatively associated with gestational age (Spearman’s rho=–0.25, p=0.0096). pTau217 was also negatively associated with birthweight (Spearman’s rho=–0.23, p=0.019) and head circumference (Spearman’s rho=–0.24, p=0.016), however, these associations were not significant when controlling for gestational age (Table S4). BD-tau did not demonstrate evidence of association with fetal parameters (Figure 2).

**Figure 2:**
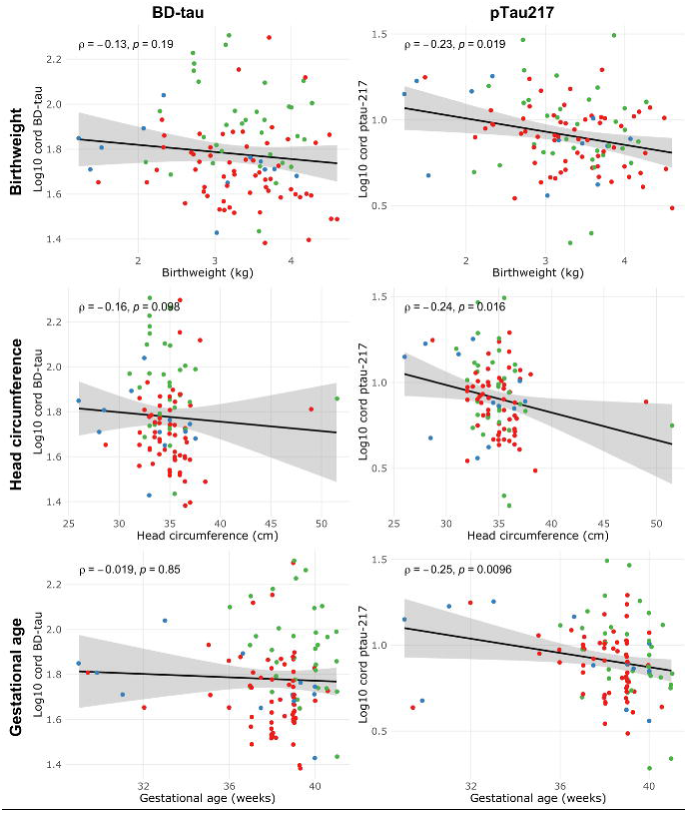
Correlation between BD-tau and pTau217 and fetal factors—birthweight, head circumference, and gestational age. Spearman correlation coefficients are shown. Red dot: elective Caesarean section (CS); blue dot: emergency CS; green dot: vaginal delivery.

##### Relationship between tau and maternal factors

A total of 8 participants were classified as having maternal hypertension—either chronic hypertension, gestational hypertension, or pre-eclampsia. Both BD-tau and pTau217 were positively associated with maternal hypertension (median BD-tau: 81.0 vs. 54.9pg/mL, Wilcoxon p=0.026; median pTau217: 10.3 vs. 7.7pg/mL, Wilcoxon p=0.029), but were not associated with other maternal health conditions (Figures S2 and S3).

##### Relationship between tau and intrapartum factors

BD-tau was associated with mode of delivery (Kruskal–Wallis p<0.001) (Figure 3), with BD-tau levels being higher with vaginal delivery compared to CS. On pairwise comparisons, no statistically significant difference was detected in BD-tau level between elective and emergency CS (median 48.4 vs. 53.6pg/mL, Wilcoxon p=0.22). pTau217 did not demonstrate a significant association with mode of delivery (Kruskal–Wallis p=.420). pTau217 was negatively associated with the duration of the second stage of labour (Spearman rho –0.4, p=0.042), whilst BD-tau and other biomarkers were not (Figure S4).

**Figure 3:**
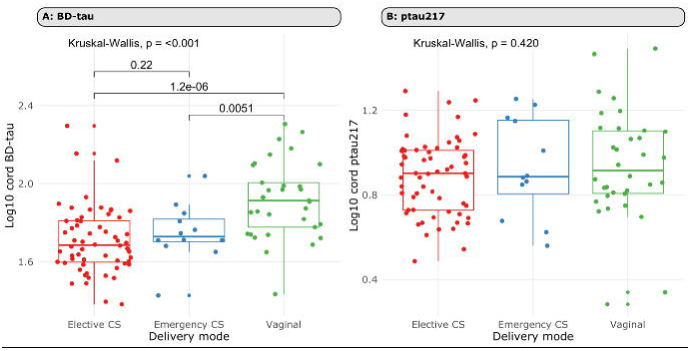
Correlation between tau and mode of delivery. A: Correlation between Log_10_ BD-tau and mode of delivery. Wilcoxon p-values are shown for comparisons between individual modes of delivery. B: Correlation between Log_10_ pTau217 and mode of delivery. Red dot: elective caesarean section (CS); blue dot: emergency CS; green dot: vaginal delivery.

### Correlation between tau and other biomarkers

Correlations between cord blood tau and other biomarkers are demonstrated in Figure 4. BD-tau demonstrated a positive association with pTau217 (Spearman’s rho=0.66, p<0.001), NfL (Spearman’s rho=0.58, p<0.001), and lactate (Spearman’s rho=0.34, p<0.001), and a negative association with cord pH (Spearman’s rho=–0.29, p=0.003) and base excess (Spearman’s rho=–0.35, p<0.001). pTau217 demonstrated a positive correlation with NfL (Spearman’s rho=0.44, p<0.001), but was not associated with other biomarkers. We did not observe evidence of a difference between pTau217 and BD-tau in the strength of the positive correlation with NfL (z=1.48, p=0.140) (Table S5).

**Figure 4:**
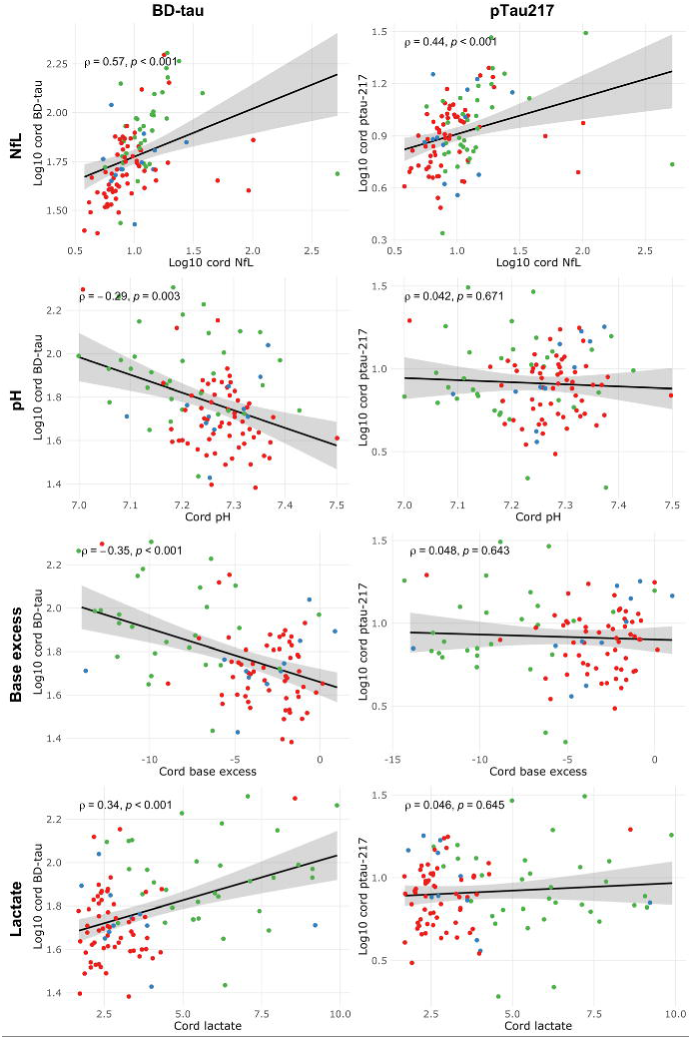
Correlation between cord blood biomarkers. Association of BD-tau with Neurofilament light chain (NfL) (n=105), pH (n=104), base excess (n=94), and lactate (n=103); association of pTau217 and NfL (n=106), pH (n=105), base excess (n=95), and lactate (n=104). Red dot: elective caesarean section (CS); blue dot: emergency CS; green dot: vaginal delivery.

### Relationship between tau and fetoplacental outcome

A total of 11 participants had reported placental abnormalities, 6 of which were defined as histopathological placental abnormalities (Appendix 1). Adjusting for low birth weight, the presence of histopathological placental abnormalities was positively associated with BD-tau and pTau217 (Table S6).

pTau217 levels were associated with preterm birth (median 7.7 in term participants vs. 12.2pg/mL in preterm participants; Wilcoxon p=0.010). In addition, pTau217 levels were positively associated with respiratory complications (median 7.6 vs. 10.4pg/mL; Wilcoxon p=0.012) and resuscitation requirement at birth (median pTau217: 7.6 vs. 10.2pg/mL; Wilcoxon p=0.027), however these associations were not observed when adjusting for preterm birth (Table S7).

## Discussion

### Main findings

Our study adds credence to the use of cord blood tau as a biomarker of neurodevelopment, perinatal hypoxia, and CNS injury. It is the first to examine associations between fetomaternal outcomes and cord blood subtypes of tau—BD-tau and pTau217. We observed a correlation between BD-tau and surrogate markers of hypoxia, including serum biomarkers of anaerobic metabolism and non-reassuring fetal status. Additionally, we found associations between tau and maternal and fetoplacental factors that could contribute to increased baseline risk of encephalopathy, including preterm birth, placental abnormalities, and maternal hypertension. Understanding the relationship between the pathogenesis of hypoxic brain injury, neuron-specific biomarkers, and clinically observed outcome could aid diagnosis, prognosis, and treatment of patients suffering from HIE.

### Clinical implications and future research

By addressing surrogate markers of fetal and neonatal hypoxia, our study of predominantly ‘healthy’ participants aims to link blood tau levels with risk factors for HIE, and provide a foundation for future research into the clinical use of cord blood tau in HIE diagnosis. Our study demonstrated a strong correlation between tau levels and non-specific markers of acidosis and anaerobic metabolism, such as neonatal pH and lactate levels, extending previous research.^1,18,26–28^ Importantly, our study is the first to examine BD-tau specifically, which offers an advantage over these traditional biomarkers due to its greater specificity for central nervous system (CNS) injury.^18,26–28^ Previous studies have mainly involved case-control analyses of HIE cohorts, demonstrating a positive correlation between early serum tau levels, HIE diagnosis based on EEG and clinical criteria, as well as HIE severity.^26,28^

The proposed role of tau as a surrogate marker of fetal hypoxia is further evidenced by the correlation between early cord blood BD-tau and non-reassuring fetal status observed in this study. This finding was not statistically significant when adjusting for mode of delivery and gestational age. However, operative delivery is commonly performed for fetal distress, and adjusting for mode of delivery could therefore cause the association between BD-tau and fetal distress to be underestimated. Fetal distress, indicated by pathological fetal heart rate abnormalities, may signal early hypoxia compensation and correlate with neonatal acidosis, low APGAR scores, NICU admission, and resuscitation.^13,29^ However, CTG interpretation is subjective, operator-dependent, and has low predictive value for long-term outcomes.^29^ Biomarkers like BD-tau could improve diagnostic specificity and guide interventions such as therapeutic hypothermia. BD-tau may offer a more specific marker of neurological damage compared current blood biomarkers.^18,26–28^

Our study also explored potential correlations between neonatal tau levels and antenatal risk factors for adverse neurological outcomes. Macroscopic (e.g., cord knots, marginal cord insertion) and microscopic (e.g., avascular villi) abnormalities in the ‘placenta–brain axis’ can contribute to intrauterine growth restriction and increased susceptibility of the fetal brain to injury during acute stress.^30,31^ We observed increased pTau217 levels in participants with histopathological placental abnormalities. Our findings should prompt further investigation into whether excessive tau phosphorylation may be related to aberrant placental function, indicating potential increased risk of neurological injury.

Microscopic placental changes may also occur as a consequence of pre-eclampsia (PET).^32^ Non-PET hypertensive disorders may show some similar microvascular changes to a lesser unknown degree, manifesting in increased neonatal stroke risk.^32^ Associations between increased tau and hypertension should be further explored to identify patients with microvascular placental changes who may be more vulnerable to physiologic stress and sentinel intrapartum events.

Phosphorylated tau (pTau217) showed a positive correlation with gestational age but not hypoxic markers. While tau phosphorylation is essential for neuronal function, excessive phosphorylation may contribute to neuronal dysfunction.^33,34^ In adults, pTau217 elevation reflects blood-brain barrier disruption and protein aggregation rather than direct neuronal injury.^33,34^ It remains unclear whether high neonatal pTau217 signals abnormal neuronal development or increased neuropathology risk. Further research is needed to clarify these mechanisms and define threshold levels for clinical significance.

### Strengths and limitations

This study is the first to distinguish between specific tau subtypes—rather than relying solely on total tau levels—paving the way for more precise diagnosis and prognosis. Correlations between BD-tau and surrogate markers of hypoxia form a strong foundation for future research into the use of cord blood tau in birth asphyxia screening. The successful sample collection and analysis, using validated assays, highlights the potential for tau cord blood measurement to feasibly be incorporated into neonatal care in an Australian context.

Limitations of this study included the relatively small sample size, from a single centre. In addition, the largely ‘healthy’ maternal cohort and short-term follow-up limited adverse outcome data. Also, our study only included static biomarker measurement at time of delivery, missing serial neonatal samples. Late serum tau rises have been linked to neurodevelopmental issues at 1–2 years,^35^ highlighting the need for further research on serial tau measurements.

## Conclusions

This study establishes BD-tau and pTau217 as potential biomarkers for fetal hypoxia and neurological vulnerability. BD-tau correlated with fetal distress, cord lactate, and pH, while phosphorylated tau levels linked to maternal hypertension and placental abnormalities. Larger, longitudinal studies are needed to validate these findings and explore the integration of tau biomarkers into neonatal screening tools.

## Data Availability

All data produced in the present study are available upon reasonable request to the authors subject to ethics approval constraints.

## Acknowledgements

HZ is a Wallenberg Scholar and a Distinguished Professor at the Swedish Research Council supported by grants from the Swedish Research Council (#2023-00356; #2022-01018 and #2019-02397), the European Union’s Horizon Europe research and innovation programme under grant agreement No 101053962, Swedish State Support for Clinical Research (#ALFGBG-71320), the Alzheimer Drug Discovery Foundation (ADDF), USA (#201809-2016862), the AD Strategic Fund and the Alzheimer’s Association (#ADSF-21-831376-C, #ADSF-21-831381-C, #ADSF-21-831377-C, and #ADSF-24-1284328-C), the European Partnership on Metrology, co-financed from the European Union’s Horizon Europe Research and Innovation Programme and by the Participating States (NEuroBioStand, #22HLT07), the Bluefield Project, Cure Alzheimer’s Fund, the Olav Thon Foundation, the Erling-Persson Family Foundation, Familjen Rönströms Stiftelse, Stiftelsen för Gamla Tjänarinnor, Hjärnfonden, Sweden (#FO2022-0270), the European Union’s Horizon 2020 research and innovation programme under the Marie Skłodowska-Curie grant agreement No 860197 (MIRIADE), the European Union Joint Programme – Neurodegenerative Disease Research (JPND2021-00694), the National Institute for Health and Care Research University College London Hospitals Biomedical Research Centre, and the UK Dementia Research Institute at UCL (UKDRI-1003).

## Contribution of Authorship

RDS designed the study in consultation with BdV, KK, BM, FG, and HM. FG, HZ and KB supplied the assays and managed biofluid analysis. KK consulted hospital records for relevant cohort data. EP and TP conducted the statistical analysis. EP, FG, and TP drafted the manuscript. All authors provided critical feedback on the manuscript.

## Ethics approval

Ethics approval was granted by the Sydney Local Health District Human Research and Ethics Committee (approval reference: 2022/ETH01100). Data were reported according to the Strengthening the Reporting of Observational Studies in Epidemiology (STROBE) guidelines.

## References

1. Gaulee P, Yang Z, Sura L, et al. Concentration of Serum Biomarkers of Brain Injury in Neonates With a Low Cord pH With or Without Mild Hypoxic-Ischemic Encephalopathy. Front Neurol. 2022;13:934755. doi:10.3389/fneur.2022.934755

2. Jacobs SE, Berg M, Hunt R, Tarnow-Mordi WO, Inder TE, Davis PG. Cooling for newborns with hypoxic ischaemic encephalopathy. Cochrane Database Syst Rev. 2013;2013(1):CD003311. doi:10.1002/14651858.CD003311.pub3

3. Leijser LM, Vein AA, Liauw L, Strauss T, Veen S, Wezel-Meijler G van. Prediction of short-term neurological outcome in full-term neonates with hypoxic-ischaemic encephalopathy based on combined use of electroencephalogram and neuro-imaging. Neuropediatrics. 2007;38(5):219–227. doi:10.1055/s-2007-992815

4. Sarnat HB, Sarnat MS. Neonatal encephalopathy following fetal distress. A clinical and electroencephalographic study. Arch Neurol. 1976;33(10):696–705. doi:10.1001/archneur.1976.00500100030012

5. Finder M, Boylan GB, Twomey D, Ahearne C, Murray DM, Hallberg B. Two-Year Neurodevelopmental Outcomes After Mild Hypoxic Ischemic Encephalopathy in the Era of Therapeutic Hypothermia. JAMA Pediatr. 2020;174(1):48. doi:10.1001/jamapediatrics.2019.4011

6. Sarnat HB, Flores-Sarnat L, Fajardo C, Leijser LM, Wusthoff C, Mohammad K. Sarnat Grading Scale for Neonatal Encephalopathy after 45 Years: An Update Proposal. Pediatr Neurol. 2020;113:75–79. doi:10.1016/j.pediatrneurol.2020.08.014

7. Fiala M, Baumert M, Surmiak P, Walencka Z, Sodowska P. Umbilical markers of perinatal hypoxia. Ginekol Pol. 2016;87(3):200–204. doi:10.17772/gp/60552

8. Gonzalez-Ortiz F, Dulewicz M, Ashton NJ, et al. Association of Serum Brain-Derived Tau With Clinical Outcome and Longitudinal Change in Patients With Severe Traumatic Brain Injury. JAMA Netw Open. 2023;6(7):e2321554. doi:10.1001/jamanetworkopen.2023.21554

9. Gundersen JK, Gonzalez-Ortiz F, Karikari T, et al. Clinical value of plasma brain-derived tau and p-tau217 in acute ischemic stroke. Cereb Circ - Cogn Behav. 2024;6:100291. doi:10.1016/j.cccb.2024.100291

10. Ashton NJ, Brum WS, Di Molfetta G, et al. Diagnostic Accuracy of a Plasma Phosphorylated Tau 217 Immunoassay for Alzheimer Disease Pathology. JAMA Neurol. Published online January 22, 2024:e235319. doi:10.1001/jamaneurol.2023.5319

11. Rubenstein R, Chang B, Yue JK, et al. Comparing Plasma Phospho Tau, Total Tau, and Phospho Tau–Total Tau Ratio as Acute and Chronic Traumatic Brain Injury Biomarkers. JAMA Neurol. 2017;74(9):1063. doi:10.1001/jamaneurol.2017.0655

12. Gonzalez-Ortiz F, Turton M, Kac PR, et al. Brain-derived tau: a novel blood-based biomarker for Alzheimer’s disease-type neurodegeneration. Brain. 2023;146(3):1152–1165. doi:10.1093/brain/awac407

13. Gonzalez-Ortiz F, Ferreira PCL, González-Escalante A, et al. A novel ultrasensitive assay for plasma p-tau217: Performance in individuals with subjective cognitive decline and early Alzheimer’s disease. Alzheimers Dement. 2024;20(2):1239–1249. doi:10.1002/alz.13525

14. Khong TY, Mooney EE, Ariel I, et al. Sampling and Definitions of Placental Lesions: Amsterdam Placental Workshop Group Consensus Statement. Arch Pathol Lab Med. 2016;140(7):698–713. doi:10.5858/arpa.2015-0225-CC

15. East CE, Leader LR, Sheehan P, Henshall NE, Colditz PB, Lau R. Intrapartum fetal scalp lactate sampling for fetal assessment in the presence of a non-reassuring fetal heart rate trace. Cochrane Database Syst Rev. 2015;2015(5):CD006174. doi:10.1002/14651858.CD006174.pub3

16. Fetal blood sampling in intrapartum care: SESLHD clinical procedure. Published online 2019.

17. New South Wales Ministry of Health. Heart rate monitoring guide. Published online 2018. https://www1.health.nsw.gov.au/pds/Pages/doc.aspx?dn=GL2018_025

18. Toorell H, Zetterberg H, Blennow K, Sävman K, Hagberg H. Increase of neuronal injury markers Tau and neurofilament light proteins in umbilical blood after intrapartum asphyxia. J Matern Fetal Neonatal Med. 2018;31(18):2468–2472. doi:10.1080/14767058.2017.1344964

19. ANZCOR (Australian and New Zealand Committee on Resuscitation). Guideline 13.1 – Introduction to Resuscitation of the Newborn. Published online 2024. https://www.anzcor.org/home/neonatal-resuscitation/guideline-13-1-introduction-to-resuscitation-of-the-newborn/

20. Zalcberg D, Kramer K, Payne E, et al. The association of umbilical cord blood neurofilament light with non-reassuring fetal status: a prospective observational study. Published online January 25, 2025. doi:10.1101/2025.01.23.25320706

21. Hernan M, Robins J. Causal Inference: What If. Boca Raton: Chapman & Hall/CRC; 2020. https://www.hsph.harvard.edu/miguel-hernan/wp-content/uploads/sites/1268/2024/01/hernanrobins_WhatIf_2jan24.pdf

22. Meng X li, Rosenthal R, Rubin DB. Comparing correlated correlation coefficients. Psychol Bull. 1992;111(1):172–175. doi:10.1037/0033-2909.111.1.172

23. World Health Organization. Low birth weight. Published online 2024. Accessed September 19, 2024. https://www.who.int/data/nutrition/nlis/info/low-birth-weight#:~:text=Low%20birth%20weight%20has%20been,2500%20grams%20(5.5%20pounds).

24. Diedenhofen B, Musch J. cocor: a comprehensive solution for the statistical comparison of correlations. PloS One. 2015;10(3):e0121945. doi:10.1371/journal.pone.0121945

25. Sachs M. plotROC: A Tool for Plotting ROC Curves. J Stat Softw Code Snippets. 2017;79(2):1–19. doi:doi:10.18637/jss.v079.c02

26. Wu H, Li Z, Yang X, Liu J, Wang W, Liu G. SBDPs and Tau proteins for diagnosis and hypothermia therapy in neonatal hypoxic ischemic encephalopathy. Exp Ther Med. 2017;13(1):225–229. doi:10.3892/etm.2016.3911

27. Chavez-Valdez R, Miller S, Spahic H, et al. Therapeutic Hypothermia Modulates the Relationships Between Indicators of Severity of Neonatal Hypoxic Ischemic Encephalopathy and Serum Biomarkers. Front Neurol. 2021;12:748150. doi:10.3389/fneur.2021.748150

28. Toorell H, Carlsson Y, Hallberg B, et al. Neuro-Specific and Immuno-Inflammatory Biomarkers in Umbilical Cord Blood in Neonatal Hypoxic-Ischemic Encephalopathy. Neonatology. Published online September 29, 2023:1–9. doi:10.1159/000533473

29. Jonsson M, Ågren J, Nordén-Lindeberg S, Ohlin A, Hanson U. Neonatal encephalopathy and the association to asphyxia in labor. Am J Obstet Gynecol. 2014;211(6):667.e1-8. doi:10.1016/j.ajog.2014.06.027

30. Penn AA, Wintermark P, Chalak LF, et al. Placental contribution to neonatal encephalopathy. Semin Fetal Neonatal Med. 2021;26(4):101276. doi:10.1016/j.siny.2021.101276

31. Fox A, Doyle E, Geary M, Hayes B. Placental pathology and neonatal encephalopathy. Int J Gynaecol Obstet Off Organ Int Fed Gynaecol Obstet. 2023;160(1):22–27. doi:10.1002/ijgo.14301

32. Mann JR, McDermott S, Pan C, Hardin JW. Maternal hypertension and intrapartum fever are associated with increased risk of ischemic stroke during infancy. Dev Med Child Neurol. 2013;55(1):58–64. doi:10.1111/j.1469-8749.2012.04409.x

33. Flavin WP, Hosseini H, Ruberti JW, Kavehpour HP, Giza CC, Prins ML. Traumatic brain injury and the pathways to cerebral tau accumulation. Front Neurol. 2023;14:1239653. doi:10.3389/fneur.2023.1239653

34. Palmqvist S, Janelidze S, Quiroz YT, et al. Discriminative Accuracy of Plasma Phospho-tau217 for Alzheimer Disease vs Other Neurodegenerative Disorders. JAMA. 2020;324(8):772–781. doi:10.1001/jama.2020.12134

35. Juul SE, Voldal E, Comstock BA, et al. Association of High-Dose Erythropoietin With Circulating Biomarkers and Neurodevelopmental Outcomes Among Neonates With Hypoxic Ischemic Encephalopathy: A Secondary Analysis of the HEAL Randomized Clinical Trial. JAMA Netw Open. 2023;6(7):e2322131. doi:10.1001/jamanetworkopen.2023.22131

