## Supplementary for "Umbilical cord blood pTau217 and BD-tau are associated with markers of neonatal hypoxia: a prospective cohort study"

**Supplementary Tables**

Table S1: Red Zone fetal heart rate patterns

Reproduced from the NSW Ministry of Health heart rate monitoring guidelines.

bpm = beats per minute

| **Baseline** | **Variability** | **Decelerations** |
| --- | --- | --- |
| <100 for >10 minutes | Reduced $\leq$ 5bpm or absent | Repetitive complicated variable |
|  | Increased >25 for >30 minutes | Repetitive late |
|  | Sinusoidal pattern >30 minutes | Single prolonged (>3 minutes and no signs recovery) |

Table S2. Demographic data of BD-tau and pTau217 cohorts. All neonates with values for BD-tau (n = 105) and pTau217 (n = 106) are included in this table (3 participants had data for one biomarker but not the other).

| **Characteristic** | **N = 107***^a^* | **Missing observations** |
| --- | --- | --- |
| **Maternal Factors** | | |
| Age | 35 (33, 39) | 0 |
| BMI | 26 (23, 30) | 6 |
| Diabetes – pre-existing or gestational | 21 (20%) | 3 |
| Anaemia | 30 (29%) | 3 |
| Smoking | 4 (3.8%) | 2 |
| Hypertension | 8 (7.5%) | 1 |
| Maternal infection | 10 (9.3%) | 0 |
| **Delivery Factors** | | |
| Gestational age (weeks) | 39.0 (37.9, 39.1) | 0 |
| Preterm birth | 11 (10%) | 1 |
| Caesarean section | 72 (67%) | 0 |
| Breech presentation | 12 (13%) | 11 |
| Instrumental delivery | 35 (34%) | 4 |
| Placental abruption | 1 (1.0%) | 4 |
| Fetal distress | 20 (19%) | 3 |
| **Perinatal Outcomes** | | |
| BD-tau (pg/mL) | 56 (45, 75) | 2 |
| pTau217 (pg/mL) | 8.0 (5.8, 10.6) | 1 |
| APGAR ≤7 (1 min) | 14 (13%) | 0 |
| APGAR ≤7 (5 min) | 2 (1.9%) | 0 |
| Fetal growth restriction | 11 (10%) | 0 |
| NICU admission | 25 (23%) | 0 |
| Special care admission | 8 (7.5%) | 0 |
| Resuscitation at birth | 21 (20%) | 1 |
| Readmission to hospital | 4 (3.8%) | 3 |
| Anaesthetic complications | 7 (6.7%) | 2 |
| Birth-Related injuries | 11 (10%) | 0 |
| Neonatal sepsis | 9 (8.4%) | 0 |
| Respiratory complications | 22 (21%) | 0 |
| Gastrointestinal complications | 15 (14%) | 0 |
| Haematological complications | 3 (2.8%) | 0 |
| *^a^* Median (IQR); n (%) | | |

Table S3: Association of BD-tau with non-reassuring fetal status. A logistic regression model is used, with G-computation to compute the risk difference (per IQR increase in log10 BD-tau).

|  | **Odds Ratio (95%CI)*^a^*** | **p-value** |
| --- | --- | --- |
| Unadjusted analysis*^b^* | | |
| Not adjusted for MOD or gestational age | 3.0 (1.6, 5.7) | 0.001 |
| Analysis adjusted for gestational age + MOD*^b^* | | |
| MOD and gestational age held at mode/mean | 1.1 (0.6, 2.1) | 0.667 |
| Estimates for each MOD, holding gestational age at mean*^b^* | | |
| Elective CS | 0.8 (0.0, Inf)*^c^* | 1.000*^c^* |
| Emergency CS | 0.2 (0.0, 1.8) | 0.155 |
| Vaginal | 1.9 (0.7, 5.0) | 0.174 |
| *^a^* Odds ratios represent likelihood of fetal distress per unit increase in log10 BD-tau, adjusted for gestational age. *^b^* Model fit statistics - Unadjusted model: Log-likelihood = -42.8; Deviance = 85.6; AIC = 89.6; BIC = 94.9; No. Obs. = 102. Adjusted model: Log-likelihood = -26.7; Deviance = 53.4; AIC = 67.4; BIC = 85.8; No. Obs. = 102; p-value from LRT comparing unadjusted vs. adjusted model fit = <0.001 *^c^* Estimate is poorly defined as there was only 1 case of non-reassuring fetal status in the elective CS group | | |

Table S4: Association of cord biomarkers concentrations with birthweight and head circumference, adjusting for gestational age.

|  | **Estimate (95%CI)*^a^*** | **p-value** |
| --- | --- | --- |
| Log10 BD-tau | | |
| Birthweight (kg) | -0.04 (-0.11, 0.03) | 0.262 |
| Head circumference (cm) | -0.00 (-0.02, 0.01) | 0.649 |
| Log10 pTau217 | | |
| Birthweight (kg) | -0.06 (-0.13, 0.02) | 0.132 |
| Head circumference (cm) | -0.01 (-0.03, 0.01) | 0.179 |
| *^a^* Estimates correspond to the increase in log10 cord biomarker concentration per 1 unit increase in birthweight/head circumference, holding gestational age at its mean. | | |

Table S5: Comparisons of the relative correlations between BD-tau and pTau217 and other cord biomarkers. Positive z-statistics imply a more strongly positive correlation between BD-tau and the cord biomarker of interest, when compared to the correlation of pTau217 and the same cord biomarker.

|  | **z-statistic*^a^*** | **p-value** |
| --- | --- | --- |
| NfL | 1.475 | 0.140 |
| pH | -5.314 | <0.001 |
| Base excess | -5.599 | <0.001 |
| Lactate | 4.995 | <0.001 |
| *^a^* Calculated using the method described by Meng, Rosenthal, and Rubin (1992), for testing overlapping correlations between two dependent groups. | | |

Table S6: Correlation between Tau and histopathological placental abnormalities, adjusted for low birth weight.

|  | **Log_10_ BD-tau^a^** | | | **Log_10_ pTau217^b^** | | |
| --- | --- | --- | --- | --- | --- | --- |
| Characteristic | *Beta* | *95% CI* | *P value* | *Beta* | *95% CI* | *P value* |
| Unadjusted analysis | | | | | | |
| Histopathological placental abnormalities | | | | | | |
| Yes | 0.18 | 0.01,0.45 | 0.033* | 0.20 | 0.03,0.38 | 0.022* |
| Adjusted for low birth weight | | | | | | |
| Histopathological placental abnormalities | | | | | | |
| Yes | 0.18 | 0.01,0.34 | 0.034* | 0.20 | 0.03,0.38 | 0.024* |

^a^ Unadjusted model: Log-likelihood = 21.8; Deviance = 3.74; AIC = -37.5; BIC = -29.7; No. Obs. = 99. Adjusted model: Log-likelihood = 21.8; Deviance = 3.74; AIC = -35.5; BIC = -25.1; No. Obs. = 99; p-value from LRT comparing unadjusted vs. adjusted model fit = 0.965

^b^ Unadjusted model: Log-likelihood = 16.0; Deviance = 4.25; AIC = -26.1; BIC = -18.3; No. Obs. = 100. Adjusted model: Log-likelihood = 16.4; Deviance = 4.21; AIC = -24.9; BIC = -14.4; No. Obs. = 100; p-value from LRT comparing unadjusted vs. adjusted model fit = 0.385

^c^ CI = Confidence Interval*^5^*

^d^ *p<0.05; **p<0.01; ***p<0.001

Table S7: Association of cord pTau217 with respiratory complications and resuscitation requirement at birth. A logistic regression model is used. G-computation is used to calculate the risk difference from the logistic regression model.

|  | **Risk difference (95%CI)*^a^*** | **p-value** |
| --- | --- | --- |
| Respiratory complications*^b^* | | |
| Unadjusted analysis | 11.2% (2.6, 19.8) | 0.010 |
| Analysis adjusted for gestational age + MOD | 4.8% (-2.6, 12.2) | 0.203 |
| Resuscitation at birth*^c^* | | |
| Unadjusted analysis | 7.6% (-1.3, 16.5) | 0.095 |
| Analysis adjusted for gestational age + MOD | 0.8% (-6.9, 8.4) | 0.843 |
| *^a^* Risk difference per increase in log10 pTau217 across its interquartile range (Q3-Q1). This value is calculated from the logistic regression model using the parametric G-formula (averaging across individual contrasts for each row of the dataset). The 95%CI is calculated using the delta method. *^b^* Model fit statistics - Unadjusted model: AIC = 103; BIC = 108; No. Obs. = 105. Adjusted model: AIC = 78.9; BIC = 86.9; No. Obs. = 105; p-value from LRT comparing unadjusted vs. adjusted model fit = <0.001 *^c^* Model fit statistics - Unadjusted model: AIC = 106; BIC = 111; No. Obs. = 104. Adjusted model: AIC = 80.3; BIC = 88.2; No. Obs. = 104; p-value from LRT comparing unadjusted vs. adjusted model fit = <0.001 | | |

**Supplementary Figures**

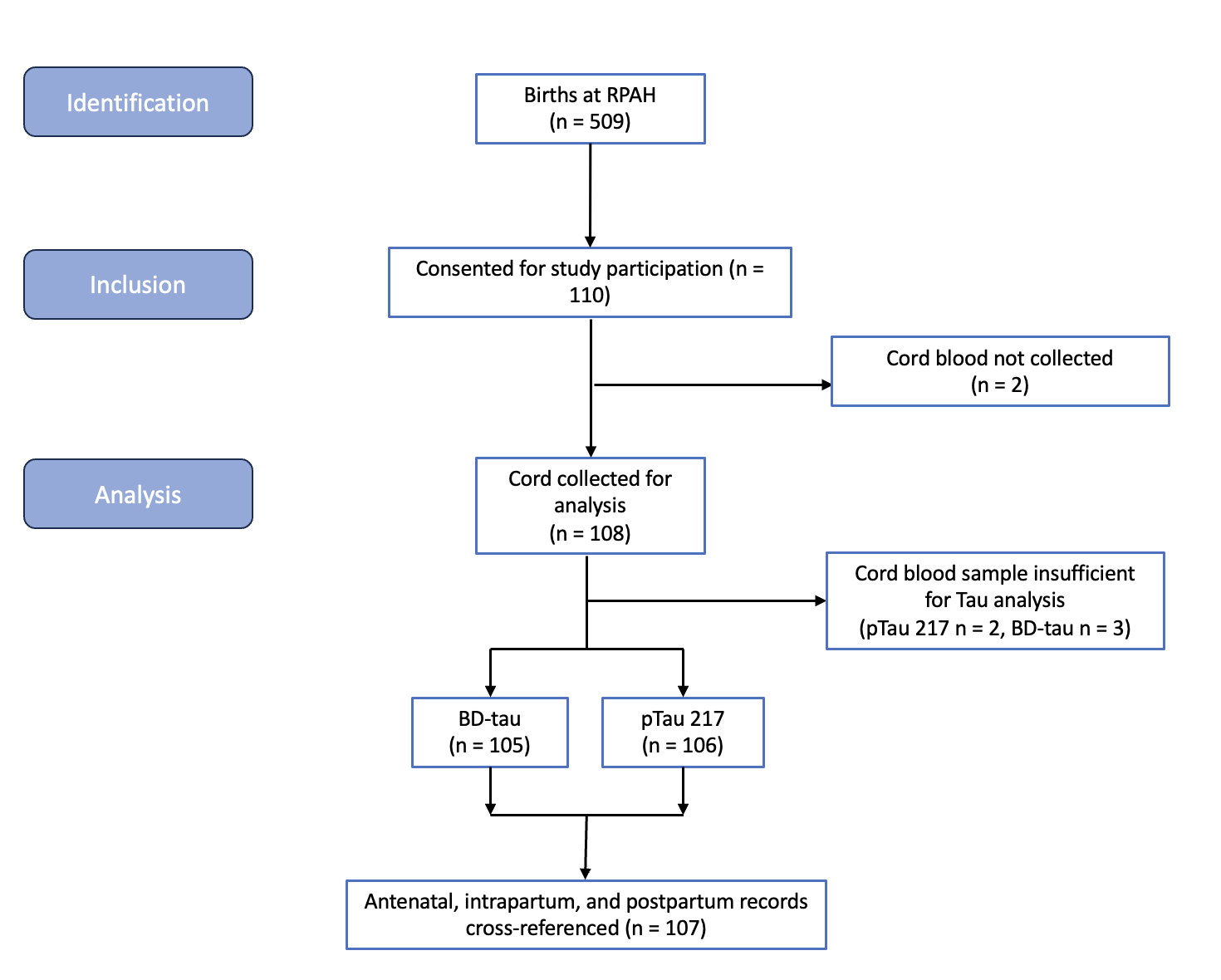

Figure S1: STROBE Diagram

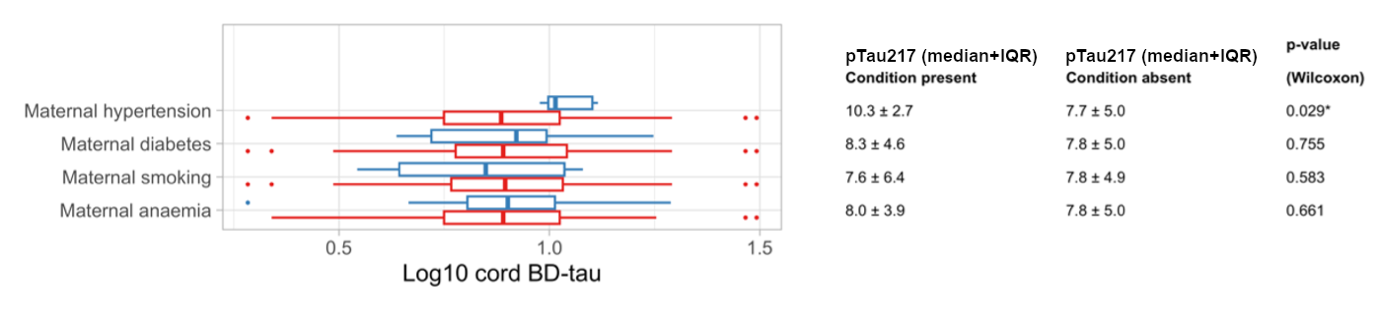

Figure S2: pTau217 levels related to maternal health conditions.

Red: condition absent; Blue: condition present.

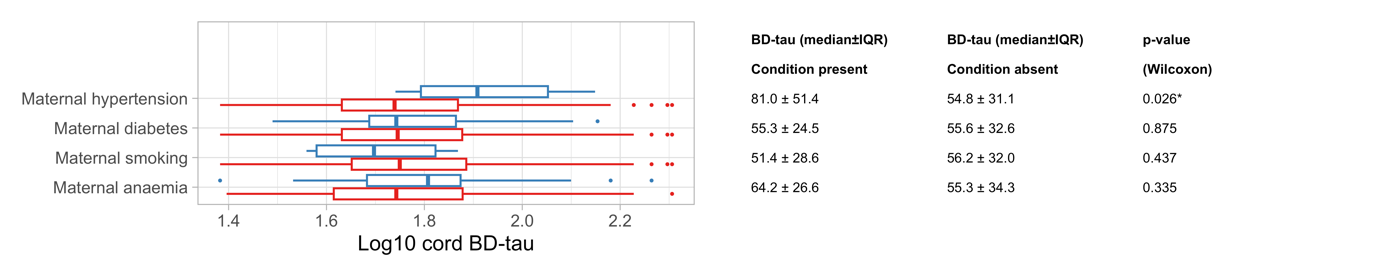

Figure S3: BD-tau levels related to maternal health conditions.

Red: condition absent; Blue: condition present.

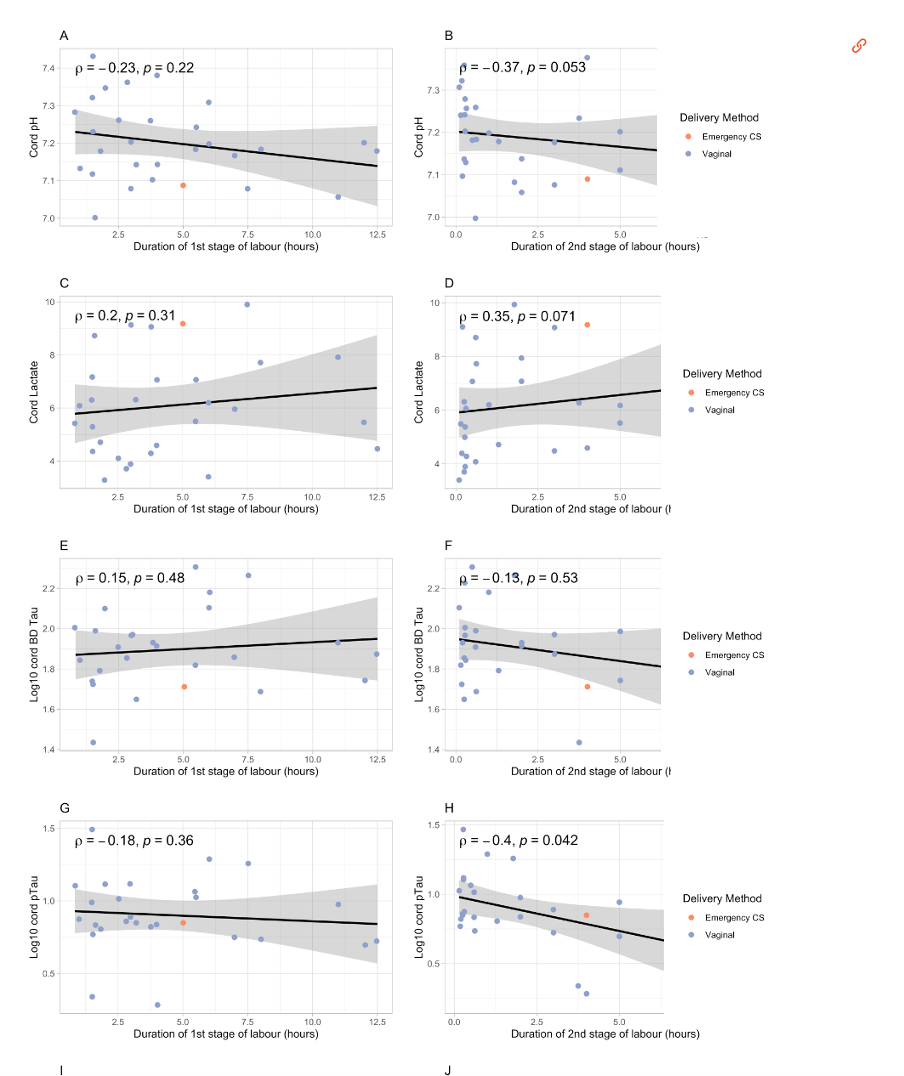

Figure S4: Association between cord blood biomarkers and duration of first and second stage of labour

(pH; lactate; log_10_ BD-tau; log_10_ pTau217). Orange denotes emergency caesarean section (Emergency CS) delivery, purple denotes vaginal delivery

Appendices

| Total | Abnormality |
| --- | --- |
| Antenatal Diagnosis | Placenta praevia |
|  | Placenta praevia |
|  | Placenta Praevia |
|  | Vasa praevia |
|  | Vasa praevia |
| Histopathological Placental Abnormality | Subchorionic haematoma with infarction |
|  | Calcification with haematoma |
|  | Calcified placenta |
|  | Succenturiate lobe |
|  | Villitis |
|  | Placental infarction |

Appendix 1: Placental abnormalities
